# Can LLMs Succeed in National Dental Examinations? A Comprehensive Analysis of Performance, Strengths, and Limitations

**DOI:** 10.64898/2026.09.30.26364385

**Authors:** Yutong Xu, Yuanyuan Peng, Dan Lv, Li Mei, Xianglong Han, Peipei Duan

## Abstract

**Background:** Large language models (LLMs) are increasingly explored as adjunct tools in graduate medical education, but their reliability for answering discipline-specific certification examinations remains incompletely characterized. In dentistry in China, the national dental examination serves as a milestone for stomatology trainees. Prior studies predominantly rely on single metric, limiting reproducibility and granularity.

**Objective:** To systematically evaluate the performance of five LLMs in answering the national standardized training examination for dental residents.

**Method:** A total of 1,318 questions covering the six dentistry disciplines were selected. Five LLMs (DeepSeek-V3, DeepSeek-R1, Qwen3-32B, Claude 3.5, and GPT-4o) were evaluated via a single-response protocol on the Dify-based Chatflow framework. Performance combined human verification with automated scoring using the Retrieval-Augmented Generation Systems (RAGAS) framework across seven dimensions. Tasks involving question classification, key-point categorization, and instructional requirement summarization were designed to enable a horizontal comparison of models’ cognitive capabilities, assessed using inter-model consistency scores.

**Results:** DeepSeek-R1 achieved the highest correct choice rate (80.90%, p < 0.001) and answer correctness score (0.853, p < 0.001), significantly outperforming the others. All models exhibited limitations, including information insufficiency, redundancy, and hallucination-related issues (lowest 44.20% in DeepSeek-R1). In classification and summarization tasks, Claude 3.5 showed higher consistency scores (186/181) than Qwen3-32B (152/151).

**Conclusion:** Evaluated LLMs demonstrated good performance on dental examination questions and in executing tasks. DeepSeek-R1 consistently outperformed the others. While LLMs showed potential as supportive tools in dental education, further refinement and validation are needed before formal adoption in graduate medical educational and clinical training settings.

## 1. Introduction

The intergration of artificial intelligence (AI) into graduate medical education (GME) has been reshaping the kandscape of intelligent healthcare, with global implications for both clinical training and competency assessment across mutiple specialties. With improvements in parameter scale and reasoning capabilities, these models have gradually transcended the boundaries of general conversation and advanced into various specialized scenarios^1^.

Existing research indicates that large language models (LLMs) demonstrate diverse potential in specialized healthcare applications and medical education. A recent comparative analysis of China’s three major medical licensing exams revealed significant disciplinary variations in ChatGPT’s performance, including its accuracy in answering questions related to clinical applications and disease management significantly exceeded that for basic medical science and pathological mechanism questions, even approaching the knowledge base of human medical students^2^. A study on Germany’s national obstetrics and gynecology exam further indicates that most answers generated by LLMs were endorsed by multiple clinical experts, suggesting potential value in developing medical education support tools^3^, however LLMs still exhibited significant performance gaps in multiple-choice questions, revealing a weakness in relying on surface pattern matching rather than deep reasoning to assess option validity^4^.

While these studies demonstrate the application potential of LLMs in certain medical scenarios, most focus on clinical disciplines like internal medicine and obstetrics/gynecology. There remains a lack of systematic evaluation of LLMs’ comprehensive capabilities in specialized fields like dentistry, which heavily rely on visual-spatial judgment and multidisciplinary integration^5,6^. These characteristics create bottlenecks for applying current general medical LLMs in dentistry: models must not only possess specialized capabilities like recognizing oral anatomical structures and imaging data, but also integrate foundational scientific knowledge with clinical information, translating it into actionable treatment plans^7,8^. However, this bottleneck points precisely to the direction of future development— the deep integration of artificial intelligence into dental education and research will inevitably drive the evolution of oral research and teaching tools toward multifunctional, multimodal, and generative capabilities^9^.

The aim of this study was to systematically evaluate the performance of five large language models (LLMs) in answering the national standardized training examination for dental residents.

## 2. Methods

### 2.1 Data Sources for Assessment

All questions used in this study were sourced from the Chinese National Standardized Training Completion Examination for Dental Residents. Questions requiring reliance on visual aids such as diagrams and standardized textual descriptions were excluded. A total of 1,318 questions were included in the study. These questions included single-choice questions (n=1,030, 78.1%), shared-stem single-choice questions (n=150, 11.4%), and case-based multiple-choice questions (n=138, 10.5%). The questions were categorized into six main disciplines: general dentistry, prosthodontics, orthodontics, oral surgery, oral medicine, and oral pathology. This classification enables analysis of the large model’s performance and variations across different disciplines. Ethical approval was not required for this study as it did not involve ethically sensitive content.

### 2.2 Evaluation Subjects

Five representative LLMs were used in this study, including:

1. DeepSeek-V3: A massive LLM employing a Mixed-Expert (MoE) architecture with 671 billion parameters^10^.
2. DeepSeek-R1: A dense model specializing in complex reasoning tasks, with approximately 20 billion parameters^11^.
3. Qwen3-32B: A dense model with strong long-context support capabilities, featuring approximately 32 billion parameters.
4. Claude 3.5: A dense model released by Anthropic, achieving breakthrough performance in reasoning, coding, and multimodal tasks.
5. GPT-4o: OpenAI’s flagship large model employing a MoE architecture, demonstrating strong general and multimodal understanding capabilities^12^.

### 2.3 Testing Methodology Based on Dify

All LLMs tests were completed within December 2025. To ensure consistency in testing conditions and certainty in results, the temperature parameter for all models was strictly set to 0, and each question was submitted to the model only once. The testing system was built on the Large Model Application Development Platform (Dify) using the Chatflow approach (not Agent mode).

Following standard LLM evaluation practices, an external knowledge base was required to provide contextual content. To directly assess LLM question-answering and reasoning capabilities, no private knowledge base was established. Therefore, the models generated background knowledge through self-retrieval.

#### 2.3.1 Generating Knowledge Prompts

Generating knowledge prompts is a method designed through specific instructions to stimulate LLMs to perform self-retrieval or leverage their internal parameterized knowledge. Its principle lies in compelling the model to first generate background knowledge relevant to the problem content upon receiving a user query, thereby simulating the rigorous process of “retrieval before reasoning”^13^. By comparing the consistency between the model’s final answer and the background knowledge it provides, we can directly determine whether the model’s conclusion is grounded in its stated evidence. This establishes a foundation for subsequent in-depth analysis of the reliability of LLM reasoning processes.

#### 2.3.2 System Prompt

All exam documents were manually uploaded to the platform by researchers and sent to respective model interfaces. A unified system prompt was designed for this purpose. This prompt first defines the model’s role as “a professional dental medicine instructor,” with the following task steps: (1) Generate knowledge prompts (i.e., independently retrieve external websites to generate relevant background knowledge); (2) Select the correct answer; (3) Provide a detailed question analysis process; (4) Classify the question type (concept differentiation, clinical diagnosis, numerical memory/calculation, treatment measures, option elimination) and knowledge point (fundamental discipline knowledge, biomechanics/materials, analysis/diagnosis, therapeutic techniques, multidisciplinary collaboration/complications) into one of five categories; and (5) Summarize the teaching requirements for the knowledge point (understand, be familiar with, master) by selecting one of three options.

#### 2.3.3 Output Format

To standardize the output of LLMs, the prompt strictly defines its structured output format, with examples provided for single-choice questions, shared-stem single-choice questions, and case analysis multiple-choice questions, ensuring the model accurately understands and adheres to complex output requirements, from generating knowledge prompts to instructional summarization.

### 2.4 RAGAS-Based Evaluation Method

To objectively and quantitatively assess the overall quality of model-generated answers, the RAGAS evaluation framework was used to measure LLMs’ outputs.

#### 2.4.1 Evaluation Metrics Description

RAGAs is a tool specifically designed to evaluate Retrieval-Augmented Generation (RAG) systems. Its core evaluation logic involves utilizing an additional large language model as an evaluator to examine the relationship among four key components: the model’s generated answer, the ground truth answer serving as the benchmark, the context relied upon during model generation, and the query posed by the user^14^. Here, “context” refers to the complete information foundation referenced by the model before generating the final answer. RAGAS employs a series of refined metrics to perform multidimensional evaluations of the model. In classic applications, these metrics each emphasize different aspects:

1. Faithfulness (FF): The core metric of the RAGAS framework, primarily examining whether the answer generated by the large model strictly originates from the context provided by the retrieval system. It aims to identify the “hallucination” issues frequently encountered by LLMs;
2. Context Recall (CR): Measures whether the context provided by the retrieval system encompasses all key factual information present in the standard answer. A low value typically indicates significant omissions in the contextual content, rendering it insufficient to support a complete and accurate response;
3. Context Precision (CP): Focuses on context content with a higher ranking and evaluates its direct relevance to the question. A low value typically indicates that preceding information contains a significant amount of irrelevant content, which may interfere with the model’s judgment in providing answers.
4. Context Relevance Level (CRL): Primarily assesses the overall closeness of the connection between the context and the question. A low value typically indicates that the context information as a whole has a high degree of redundancy and insufficient density of effective information.
5. Answer Correctness (AC) and Answer Similarity (AS): Jointly evaluate the factual and semantic alignment between the model’s final output and the standard answer. Low values typically indicate factual errors or significant semantic discrepancies between the generated answer and the standard answer.
6. Answer Relevance (AR): Primarily determines whether the model’s response directly addresses the question. A low score typically indicates the generated answer is irrelevant or avoids the core issue.

#### 2.4.2 Evaluation Data Notes

The data sources for this RAGAS evaluation are as follows: problem description, context, and AI reply all originate from corresponding content generated by large language models. Here, the context correspond to generating knowledge prompts, while the ground truth were manually input by human evaluators. For the RAGAS evaluation metrics, uniform pass thresholds were established: FF, CR, CP, and CRL at 0.8, while AC and AS at 0.9. Scores exceeding these thresholds indicate that the large language model achieved a high score for that metric.

In addition to the RAGAS metric, the foundational indicator of Correct Choice Rate (CC) was also used to assess whether the model’s output option perfectly matches the standard answer (correct=1, incorrect=0), providing the most direct result determination and complement the answer correctness (AC) results.

It is important to note that this study primarily focuses on multiple-choice questions, where model outputs are strictly confined to the given options. The probability of severe “off-topic” responses is extremely low. Consequently, the answer relevance metric (AR), which measures the relevance of answers to questions, has limited diagnostic value in this study. Therefore, no strict pass threshold was set for AR, and it is used solely as an auxiliary reference.

#### 2.4.3 Operational Process

The specific operational process is as follows: After collecting all models’ generated responses to the 1318 test questions, each question’s “query” is used as the query, the model’s provided answer as the generated answer, and the model’s background knowledge as the context. These are then uploaded to the RAGAS system alongside the ground truth answers. Subsequently, the system automatically scores each model individually based on the aforementioned metrics, enabling comparative analysis of different LLM performances.

### 2.5 LLM Consistency Analysis

The prompts in this study predefined fixed sets of options for “question classification,” “exam point classification,” and “teaching requirement summarization.” Consequently, the models’ outputs across these three tasks can be interpreted as selections from given options. To comprehensively evaluate the answer aggregation levels of different large language models in these selective tasks, we conducted simple manual proofreading scoring and consistency statistical analysis.

The frequency and distribution of each model’s selections for the given options across the three tasks (“Question Classification,” “Exam Point Classification,” and “Teaching Requirement Summary”) were quantified to measure the consensus among the large models on each task. Following the verification rule (“If at least three of the five large models provide consistent classification answers for the same question, that classification is deemed correct, with the corresponding models scoring 1 point and inconsistent models scoring 0 points; if no three or more models agree, all models score 0 points for that question”), each question and tallied model scores were manually scored. This directly assessed classification consistency and classification capability among models based on their scores.

### 2.6 IRB Approval

Not applicable. This study did not involve human participants or animals.

## 3. Results

### 3.1 Overall Performance Across Models

The overall correct choice (CC) rates and RAGAS evaluation metrics for the five large language models (LLMs) across six dental disciplines are summarized in Fig.1. All datasets met assumptions of normality and homogeneity of variance; therefore, one-way ANOVA was applied. Significant inter-model differences were observed for both CC and Answer Correctness (AC) scores (p < 0.01).

**Figure 1.**
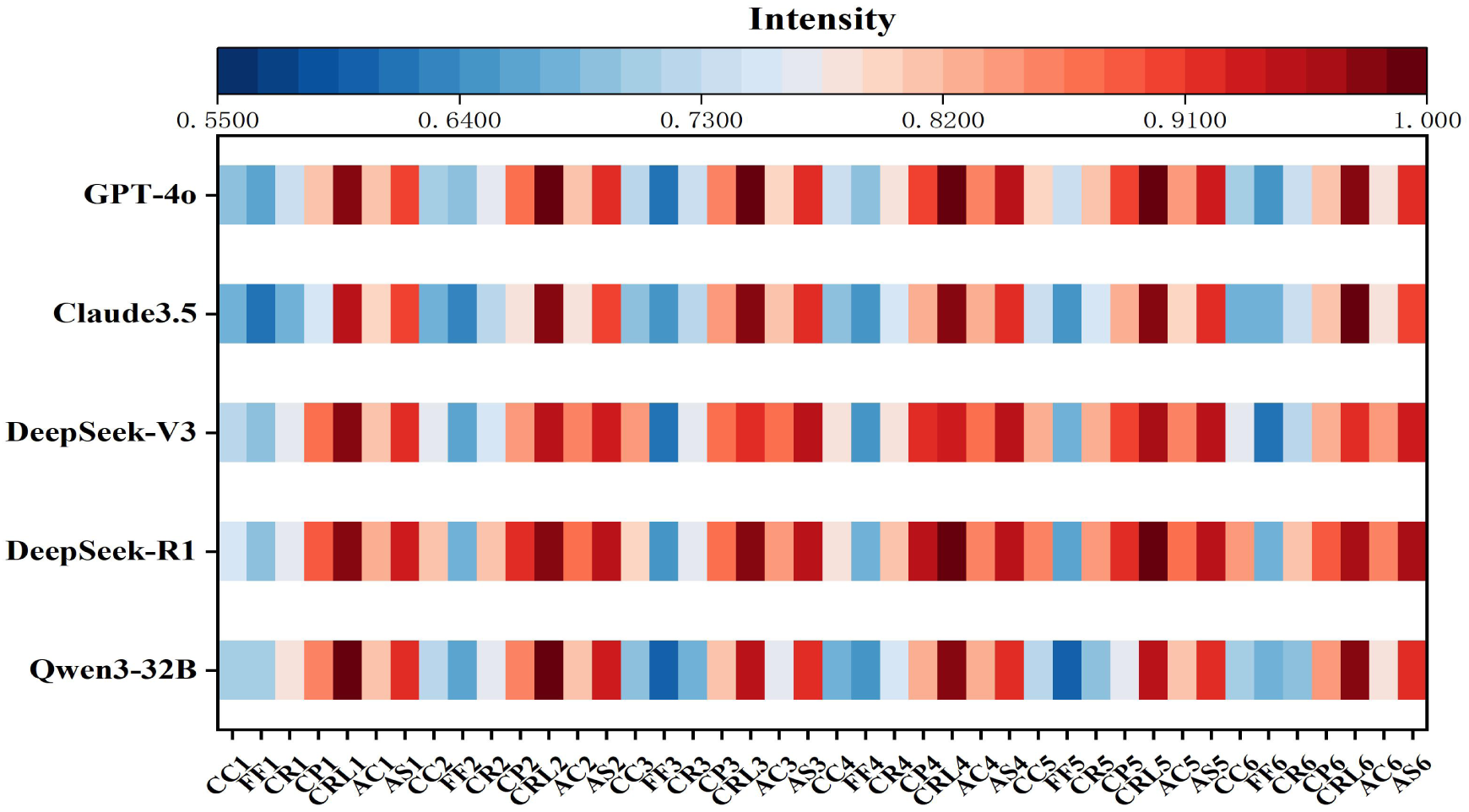
Heatmap of CC and RAGAS Metric Scores for All Large Language Models.

Across all question sets, DeepSeek-R1 achieved the highest mean accuracy (80.9%), marginally outperforming DeepSeek-V3 (78.8%). Qwen3-32B (72.5%) ranked above Claude 3.5 (70.8%) and GPT-4o (67.0%). A similar ranking was observed for AC scores, with DeepSeek-R1 (0.853) and DeepSeek-V3 (0.848) significantly outperforming the remaining models.

Using predefined RAGAS thresholds (FF, CR, CP, CRL ≥ 0.8; AC, AS ≥ 0.9), no model met the average Faithfulness (FF) threshold, and only DeepSeek-R1 satisfied the Context Recall (CR) criterion. These findings suggest persistent limitations in self-retrieval reliability across current LLMs.

### 3.2 Performance by Question Type

All five models demonstrated significantly higher accuracy on single-choice questions than on multiple-choice questions (p < 0.01; Fig.2). For single-choice items, accuracy ranged from 74.2% to 84.5%, indicating relatively stable performance. In contrast, multiple-choice accuracy varied widely (26.5%–49.9%), reflecting greater task complexity and inter-model variability. DeepSeek-R1 and DeepSeek-V3 consistently outperformed other models on multiple-choice questions, each exceeding 40% accuracy (p < 0.01).

**Figure 2.**
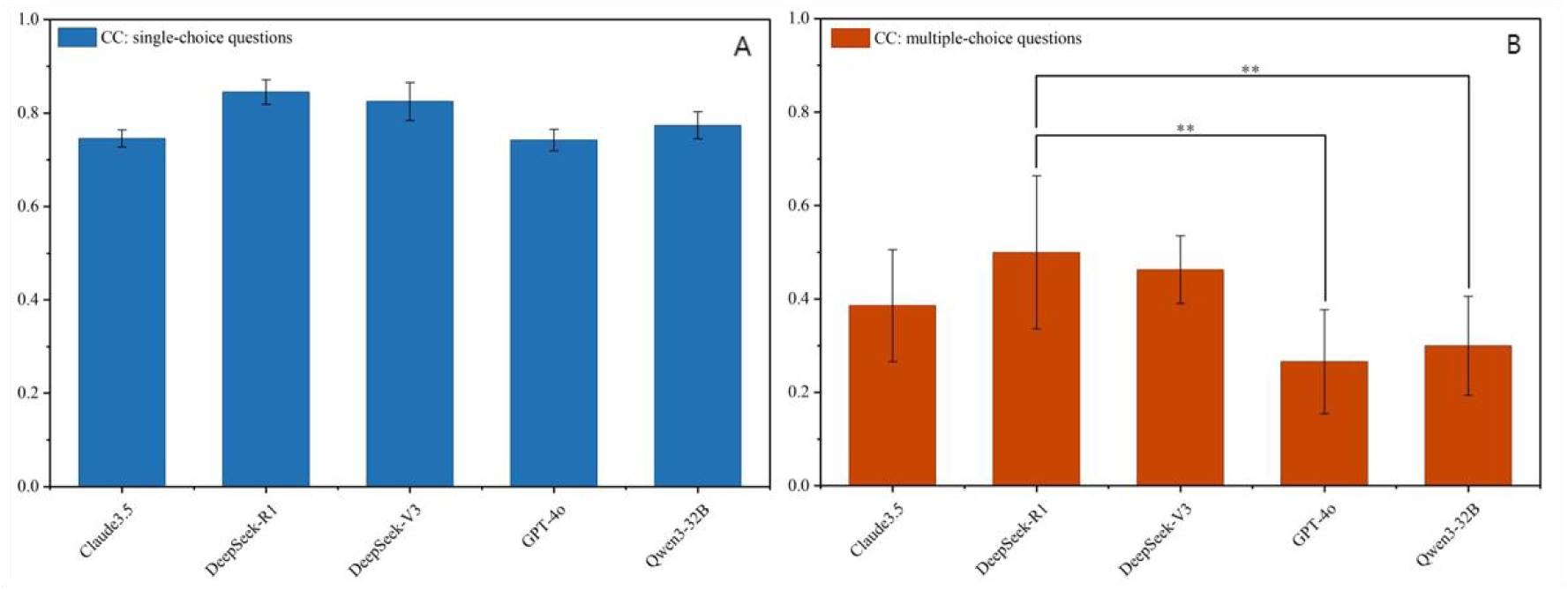
Bar Charts of CC Scores for all five models in Single-Choice and Multiple-Choice Questions.

### 3.3 Reasoning Quality and Hallucination Analysis

The relationship between Faithfulness (FF) and Answer Correctness (AC) revealed variability in reasoning reliability across models. Low FF cases—indicative of hallucination risk—were common across all models. As shown in Figure 3, DeepSeek-R1 (44.2%) and Qwen3-32B (45.5%) exhibited the lowest proportions of low-FF outputs, whereas GPT-4o showed the highest (48.3%), including the greatest share of low FF/low AC cases (26.0%).

**Figure 3.**
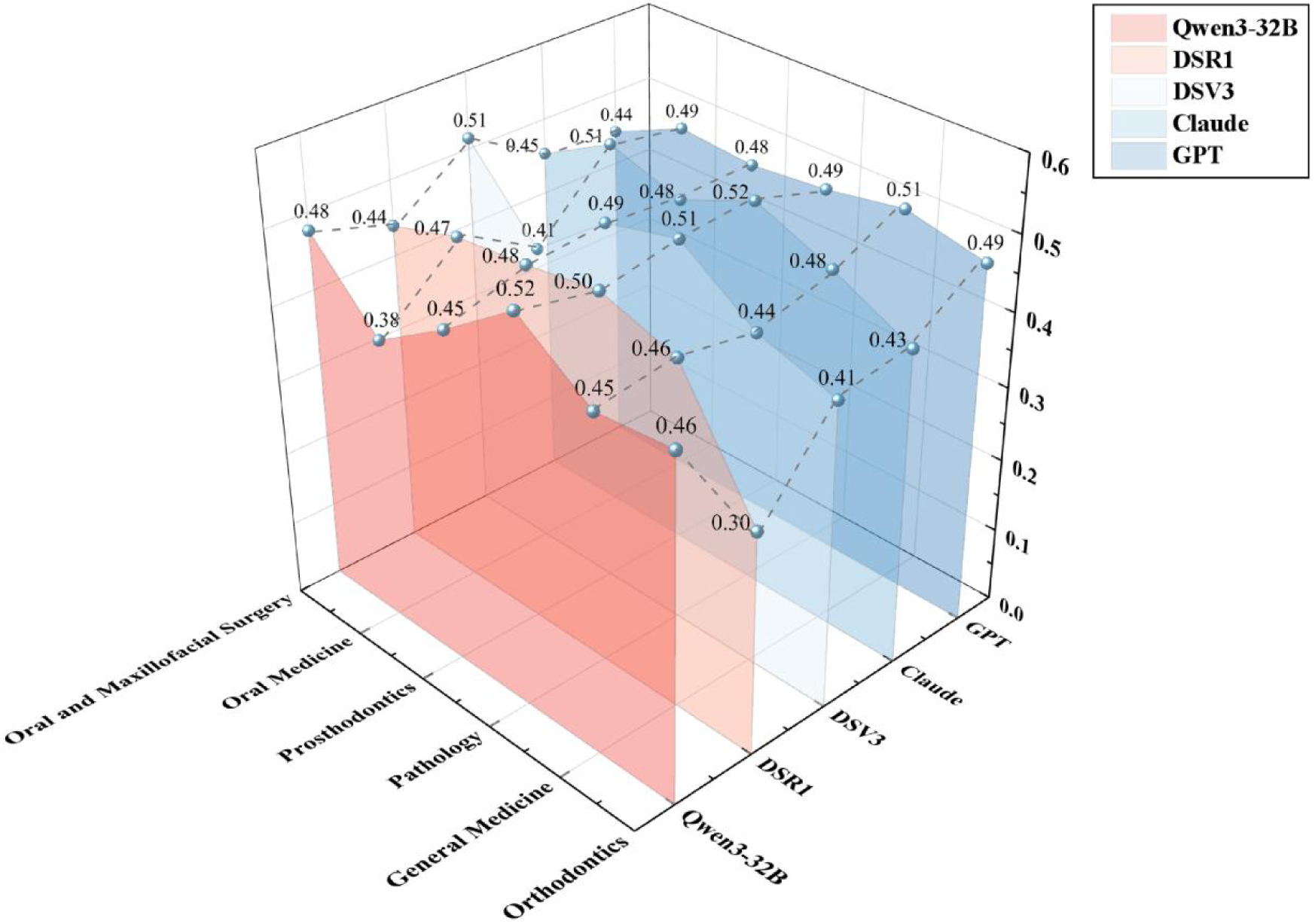
The Proportion of Questions in Each Set where the Five Large Models Achieved Low FF Scores.

A five-category error taxonomy was applied to classify reasoning failures (Fig.4). Among an average of 1,196 valid classifications per dataset, Thought Hallucination was the most frequent error type (515 instances), followed by Information Deficiency (273) and Information Noise (221). Knowledge Bias (147) and Logic Bias (39) were comparatively infrequent (p < 0.01). GPT-4o exhibited the highest frequency of Information Deficiency, while DeepSeek-R1 basically showed the lowest error counts across categories.

**Figure 4.**
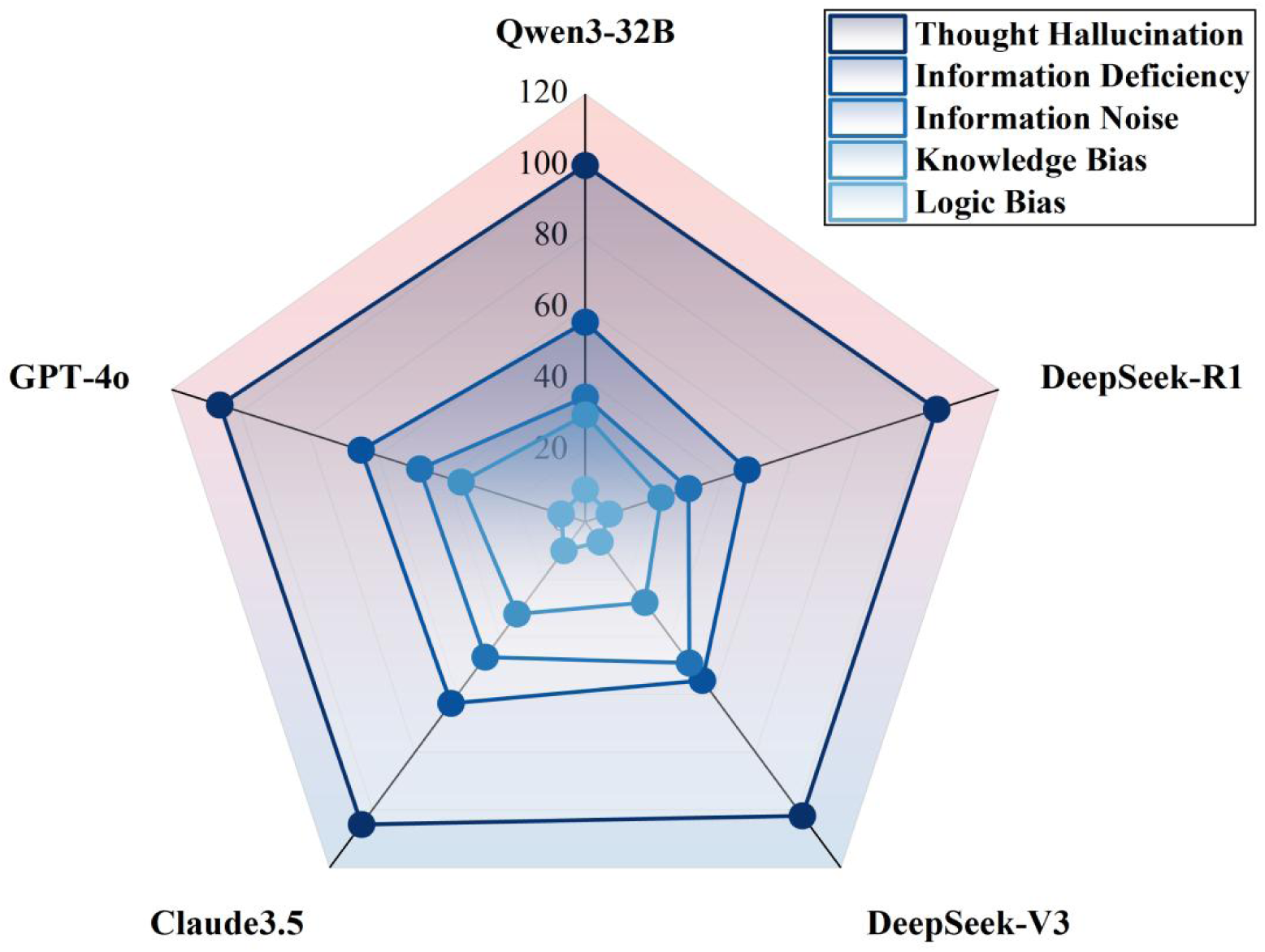
The Average Occurrence Frequency of Five Error Types across Six Sets of Questions for Each of the Five Large Models.

Discipline-specific analysis revealed notable differences: orthodontics demonstrated the lowest rate of Thought Hallucination but the highest Logic Bias, whereas oral pathology showed significantly fewer Knowledge Bias errors (p < 0.01).

### 3.4 Classification and Teaching Requirement Consistency

Across 6,590 classification selections, inter-model consistency varied substantially (p < 0.01; Fig.5). Claude 3.5 achieved the highest consistency scores in both question-type (186) and knowledge-point classification (181), indicating strong stability in labeling tasks. DeepSeek-R1 ranked second overall but showed weaker performance in maxillofacial surgery classifications. Qwen3-32B and GPT-4o demonstrated moderate but relatively uniform performance, whereas DeepSeek-V3 exhibited greater variability across disciplines.

**Figure 5.**
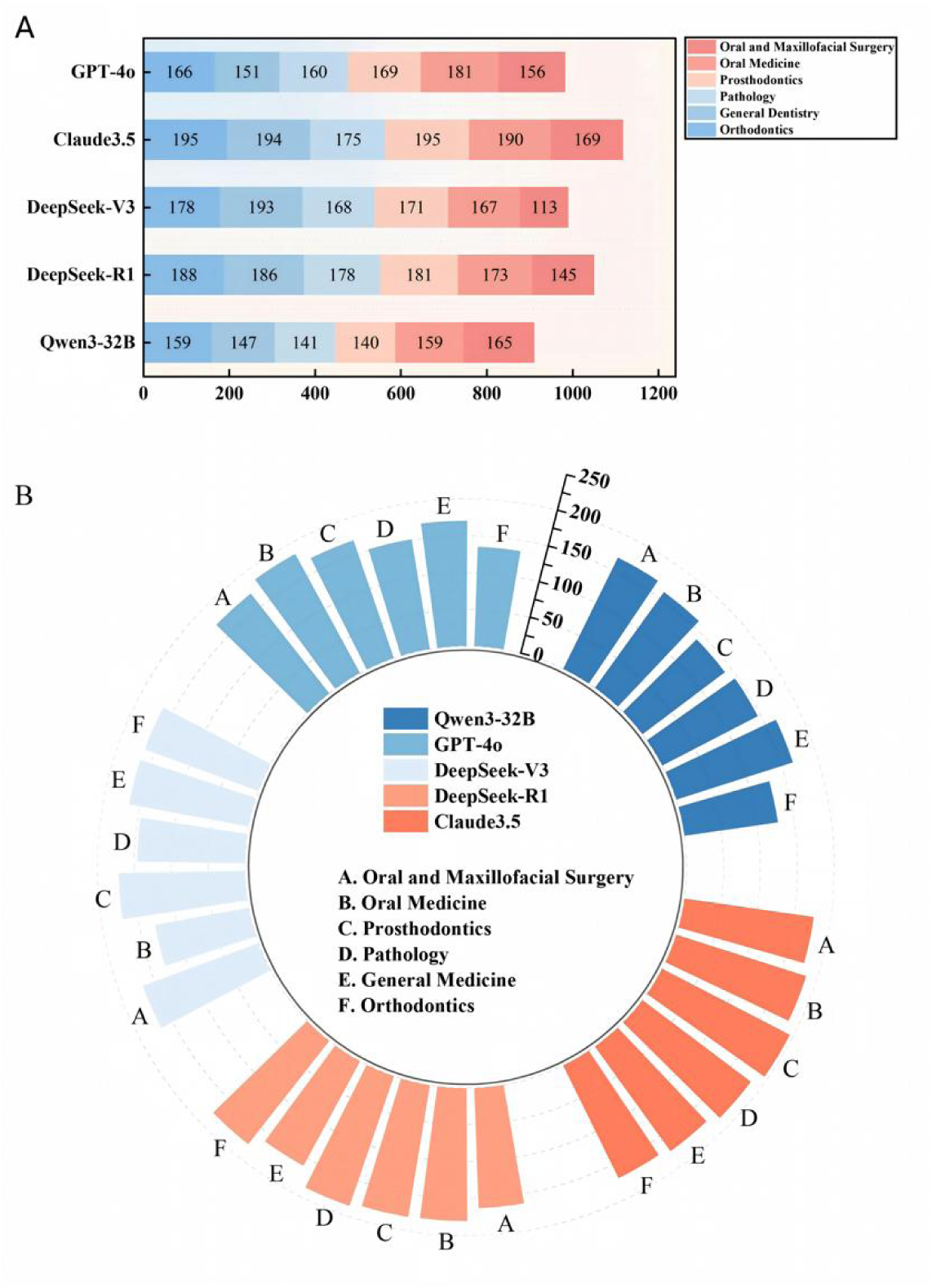
(A) Five Large Language Models Achieved Consistent Scores across Six Sets of Questions in Terms of Question Type Classification, (B) Five Large Language Models Scored Consistently across Six Sets of Questions in Terms of Knowledge Point Assessment.

For teaching requirement summarization, all models favored the “Mastery” level (57.1%), with comparatively fewer selections for “Familiarity” (32.2%) and “Understanding” (7.7%). GPT-4o showed marked divergence by selecting “Understanding” only once.

## 4. Discussion

This study provides a comprehensive evaluation of five contemporary LLMs on a national standardized dental residency examination, integrating traditional accuracy metrics with RAGAS-based reasoning quality assessment. Overall, DeepSeek-R1 demonstrated the strongest performance, achieving the highest accuracy and the most reliable reasoning outputs, suggesting meaningful potential for application in dental education support.

To effectively optimize models, this study further dissects common problem types and their occurrence patterns in large language models. Test data reveals that issues categorized as thought hallucinations, information deficiency, and information noise each appeared over 200 times on average across six test sets. Hallucinations were the most prevalent (515 instances), while knowledge bias and logic bias issues were relatively less frequent (147 and 39 instances, respectively). Independent analysis of different models reveals that all five large models averaged over 50 instances of information deficiency issues, with GPT-4o (65) and Claude 3.5 (63) exceeding 60 instances. Information noise generally ranged between 30 and 50 occurrences. While DeepSeek-V3 performed better in information deficiency (55 occurrences), it exhibited 49 information noise problems, suggesting significant redundancy in its retrieval process compared to other LLMs. Additionally, the number of knowledge bias and logic bias issues showed relatively minor differences. GPT-4o and Claude3.5 exhibited slightly more knowledge bias issues than DeepSeek-R1, DeepSeek-V3, and Qwen3-32B. This discrepancy may stem from the use of Chinese-language test questions rather than English-language ones, which could hinder the reading comprehension capabilities of foreign LLMs^15–17^. Overall, these results indicate that all large models exhibit significant issues when retrieving external public data. Their retrieved information frequently lacks critical details, contains redundant or prematurely presented irrelevant information, and may lead to fabricated reasoning or reliance on fictional knowledge—factors consistent with the high incidence of LLM hallucination-type problems^18,19^. In fact, unlike traditional models that prioritize result accuracy, medical models should focus more on the integrity of the reasoning process. If the reasoning process is ambiguous or erroneous, it may generate conclusions that appear plausible yet are erroneous; even a correct answer poses serious potential risks in practical applications and teaching^20,21^.

Improving the quality of information retrieval in large language models is a critical prerequisite for reducing model hallucinations and ensuring reliable reasoning. Based on the data from this study, we believe establishing a private domain knowledge base for oral medicine would significantly help address information retrieval deficiency in large models and facilitate the development of domain-specific LLMs for oral medicine^22^. Regarding information noise issues, the most direct improvement method is to refine the internal retrieval process of large models. This involves adding a re-ranking mechanism during information retrieval, where a cross-Encoder performs secondary scoring on the Top-K retrieval results^23^. This ensures only the most relevant and valuable content is retained. Furthermore, the lowest proportion of hallucination-related issues observed across the five large models in this evaluation was 44.197%. This rate remains excessively high for practical medical scenarios. Therefore, for applications involving large models assisting clinical diagnosis and student education, we recommend human verification of responses or using integration of AI output detectors to ensure safety and accuracy^24^.

Based on all the above evaluations, the lightweight model DeepSeek-R1 demonstrated outstanding performance over other medium or large models in the choice-based oral clinical knowledge assessments, largely consistent with the findings of TSENG C-Y and Tordjman M et al^25,26^.We attribute this advantage to DeepSeek-R1’s unique training methodology, which leverages RLVR technology, cold-start data and iterative RL fine-tuning to substantially enhance the model’s capacity for progressive analysis and logically coherent deep reasoning^27^. This enables the model to more accurately apply domain knowledge, generating clear, rigorous responses that align with medical thought processes. Notably, recent research indicates that efficient, lightweight medical vertical models built upon DeepSeek-R1 have been largely realized, effectively advancing the transformation of general-purpose large models into specialized medical tools. This model achieved a high accuracy rate of 92.1% on the USMLE, significantly outperforming other mainstream models during the same period and demonstrating robust professional problem-solving capabilities^28^.

In addition to answer accuracy, we evaluated the classification and teaching-requirement summarization capabilities of the five large language models using human-validated consistency scores. Notable inter-model differences were observed. Claude 3.5 achieved the highest overall consistency, scoring 186 in question-type classification and 181 in exam-point classification. It was the only model to exceed 180 points in both tasks, indicating strong classification stability and broad agreement with other models.

In contrast, DeepSeek-V3 demonstrated lower and more variable consistency across tasks (165 and 157), suggesting either preferential sensitivity to specific question types and knowledge domains or a distinct classification reasoning strategy. DeepSeek-R1 exhibited mixed performance, outperforming Claude 3.5 in pathology-related question and exam-point classification, but performing significantly worse in maxillofacial surgery, highlighting limitations in handling this specialty. Qwen3-32B (152, 151) and GPT-4o (164, 164) showed nearly identical scores across both classification tasks, reflecting stable but comparatively weaker agreement with the broader model group and indicating limitations in classification reliability.

Analysis of teaching-requirement label distributions revealed a generally consistent trend across models. DeepSeek-R1 and DeepSeek-V3 displayed nearly identical selection patterns, while minor discrepancies were observed between Claude 3.5 and Qwen3-32B. Notably, GPT-4o selected the “Understanding” level only once, in sharp contrast to Qwen3-32B’s 233 selections. Conversely, Claude 3.5 overwhelmingly favored the “Mastery” level, selecting it more than 900 times—substantially exceeding GPT-4o’s 595 selections. These findings indicate substantial inter-model variability in pedagogical interpretation and summarization preferences.

Overall, the results demonstrate that LLMs differ in classification consistency and summarization behavior. While individual models show strengths in specific domains, none consistently performs optimally across all classification and instructional tasks. Therefore, task-specific model selection or ensemble strategies with manually assisted may better leverage complementary model strengths and improve robustness in educational applications^29^.

There are limitations in this study. First, due to technical and resource constraints, questions requiring dental imaging interpretation were excluded, despite the central role of radiographic analysis in dental education and clinical practice. As multimodal LLM capabilities continue to advance, future evaluations should incorporate image-based assessments. Second, the absence of a domain-specific private dental knowledge base likely constrained model performance on highly specialized terminology and concepts. Integrating curated professional knowledge bases may substantially improve retrieval accuracy and reasoning reliability. Finally, given the rapid evolution of LLMs, the evaluated versions may not reflect their current peak performance^30^. The findings provide methodological guidance for future system development. Future work should prioritize the integration of dentistry-specific knowledge, the optimization of retrieval pipelines, and the development of rigorous multimodal evaluation frameworks to ensure the safe and effective deployment of LLMs in dental education and clinical decision support.

## 5. Conclusion

The five evaluated large language models demonstrated overall strong performance in responding to dental examination questions and in performing classification and summarization tasks. Among them, DeepSeek-R1 consistently achieved superior results across multiple evaluation metrics, indicating comparatively stronger reasoning capabilities and higher response accuracy. The limitations observed across all models underscore the necessity for continued methodological refinement and rigorous validation to ensure reliability, safety, and generalizability prior to their widespread integration into formal dental education and clinical training environments.

## Data Availability

All data produced in the present study are available upon reasonable request to the authors

## Declaration

### Ethical Approval

Not applicable. This study did not involve human participants or animals.

### Informed Consent

Not applicable.This study did not involve human participants.

### Statement Regarding Research Involving Human Participants and/or Animals

Not applicable.This study did not involve human participants or animals.

### Consent to Participate

Not applicable.This study did not involve human participants.

### Consent to Publish

Not applicable.This study did not involve human participants whose data would be published.

### Author’s Contribution

All authors contributed to the study conception and design.All authors read and approved the final manuscript.

### Availabilitydata and materials

Data for study available upon reasonable request.

## References

1. Rodrigues-Pereira P, Dias-Calças MAP, Moreira Mélo A, et al. Generative Artificial Intelligence-Driven Clinical Case Simulation in Temporomandibular Disorder Education: ChatGPT Versus Real Patients. J Dent Educ. Nov 12 2025;doi:10.1002/jdd.70104

2. Zong H, Li J, Wu E, Wu R, Lu J, Shen B. Performance of ChatGPT on Chinese national medical licensing examinations: a five-year examination evaluation study for physicians, pharmacists and nurses. BMC Med Educ. Feb 14 2024;24(1):143. doi:10.1186/s12909-024-05125-7

3. Riedel M, Kaefinger K, Stuehrenberg A, et al. ChatGPT’s performance in German OB/GYN exams - paving the way for AI-enhanced medical education and clinical practice. Front Med (Lausanne). 2023;10:1296615. doi:10.3389/fmed.2023.1296615

4. Hoch CC, Wollenberg B, Lüers JC, et al. ChatGPT’s quiz skills in different otolaryngology subspecialties: an analysis of 2576 single-choice and multiple-choice board certification preparation questions. Eur Arch Otorhinolaryngol. Sep 2023;280(9):4271–4278. doi:10.1007/s00405-023-08051-4

5. Patil S, Albogami S, Hosmani J, et al. Artificial Intelligence in the Diagnosis of Oral Diseases: Applications and Pitfalls. Diagnostics (Basel). Apr 19 2022;12(5)doi:10.3390/diagnostics12051029

6. Scully C, Miller CS, Aguirre Urizar JM, et al. Oral medicine (stomatology) across the globe: birth, growth, and future. Oral Surg Oral Med Oral Pathol Oral Radiol. Feb 2016;121(2):149–157.e5. doi:10.1016/j.oooo.2015.10.009

7. Gao S, Wang X, Xia Z, Zhang H, Yu J, Yang F. Artificial Intelligence in Dentistry: A Narrative Review of Diagnostic and Therapeutic Applications. Medical Science Monitor: International Medical Journal of Experimental and Clinical Research. 2025;31

8. Li Q, Li S, Fu D, et al. The Role of Emerging Digital Technologies in Revolutionizing Dental Education: A Bibliometric Analysis. J Dent Educ. Sep 3 2025;doi:10.1002/jdd.70033

9. Lee G, Yun M, Zhai X, Crippen K. Artificial Intelligence in Science Education Research: Current States and Challenges. Journal of Science Education and Technology. 2026/02/01 2026;35(1):110–127. doi:10.1007/s10956-025-10239-8

10. DeepSeek-AI, Liu A, Feng B, et al. DeepSeek-V3 Technical Report. 2024:

11. DeepSeek-AI, Guo D, Yang D, et al. DeepSeek-R1 incentivizes reasoning in LLMs through reinforcement learning. Nature. 2025;645:633–638.

12. Hurst OA, Lerer A, Goucher AP, et al. GPT-4o System Card. 2024:

13. Liu J, Liu A, Lu X, et al. Generated Knowledge Prompting for Commonsense Reasoning. 2021:

14. Shahul E, James J, Anke LE, Schockaert S. RAGAs: Automated Evaluation of Retrieval Augmented Generation. ArXiv. 2023;abs/2309.15217

15. Aidynkyzy A, Dikenelli O, Alatli O, Bora Ş. Relation Extraction Capabilities of LLMs on Clinical Text: A Bilingual Evaluation for English and Turkish. ArXiv. 2026;abs/2601.09367

16. Liu M, Okuhara T, Chang X, et al. Performance of ChatGPT Across Different Versions in Medical Licensing Examinations Worldwide: Systematic Review and Meta-Analysis. J Med Internet Res. Jul 25 2024;26:e60807. doi:10.2196/60807

17. Yacobson E, Schleifer Y, Bar-Dov Z, Rap S, Blonder R, Alexandron G. Benchmarking AI on Standard Chemistry Exams: LLMs Still Underperform Compared to High School Students. Journal of Science Education and Technology. 2026/03/28 2026;doi:10.1007/s10956-026-10310-y

18. Ebrahimzadeh A, Salili SM. Not All Needles Are Found: How Fact Distribution and Don’t Make It Up Prompts Shape Literal Extraction, Logical Inference, and Hallucination Risks in Long-Context LLMs. ArXiv. 2026;abs/2601.02023

19. Qi S, Cao R, He Y, Yuan Z. Evaluating LLMs’ Assessment of Mixed-Context Hallucination Through the Lens of Summarization. 2025:

20. Wiest IC, Bhat M, Clusmann J, Schneider CV, Jiang X, Kather JN. Large language models for clinical decision support in gastroenterology and hepatology. Nat Rev Gastroenterol Hepatol. Nov 2025;22(11):773–787. doi:10.1038/s41575-025-01108-1

21. Xu X, Chen Y, Miao J. Opportunities, challenges, and future directions of large language models, including ChatGPT in medical education: a systematic scoping review. J Educ Eval Health Prof. 2024;21:6. doi:10.3352/jeehp.2024.21.6

22. Jang D, Shangguan Z, Tegtmeyer K, et al. MedTutor: A Retrieval-Augmented LLM System for Case-Based Medical Education. 2026:

23. Granata FM, Poggi F, Mongiovì M. Enhancing Retrieval-Augmented Generation with Entity Linking for Educational Platforms. ArXiv. 2025;abs/2512.05967

24. Arantes J. Understanding Intersections Between GenAI and Pre-Service Teacher Education: What Do We Need to Understand About the Changing Face of Truth in Science Education? Journal of Science Education and Technology. 2025/12/01 2025;34(6):1311–1322. doi:10.1007/s10956-024-10189-7

25. Tordjman M, Liu Z, Yuce M, et al. Comparative benchmarking of the DeepSeek large language model on medical tasks and clinical reasoning. Nat Med. Aug 2025;31(8):2550–2555. doi:10.1038/s41591-025-03726-3

26. Tseng C-Y, Zhang D, Wang T, et al. 47B Mixture-of-Experts Beats 671B Dense Models on Chinese Medical Examinations. ArXiv. 2025;abs/2511.21701

27. Gu B, Zhou H, Segal BM, et al. Clinical-R1: Empowering Large Language Models for Faithful and Comprehensive Reasoning with Clinical Objective Relative Policy Optimization. ArXiv. 2025;abs/2512.00601

28. Zhang M, Qin J. A Method for the Architecture of a Medical Vertical Large Language Model Based on Deepseek R1. ArXiv. 2025;abs/2505.00025

29. Wang L, Li J, Zhuang B, et al. Accuracy of Large Language Models When Answering Clinical Research Questions: Systematic Review and Network Meta-Analysis. J Med Internet Res. Apr 30 2025;27:e64486. doi:10.2196/64486

30. Calvo-Utrilla M, Paños E, Ruíz-Gallardo J-R. Chatbots in Science Education: A Scoping Review of Early Empirical Evidence. Journal of Science Education and Technology. 2025/10/10 2025;doi:10.1007/s10956-025-10260-x

